# *Bifidobacterium animalis subsp. lactis* BL-11 Promotes Height Growth in Children Aged 3-7 Years: A Self-controlled Cohort Study

**DOI:** 10.1101/2024.10.21.24315871

**Authors:** Qian Wang, Yanan Yang, Yin Xiao, Ziyi Liu, Mingxi Li, Yue Yu, Nannan Wang, Junyan Li, Xiaohui Li, Chuanhui Xu, Deyun Liu, Chongming Wu

## Abstract

**Background:** Short status of children has captured extensive attention since it reflects the health status of children and affects physical and psychological health throughout children’s life. Compelling evidences have revealed that gut microbiota participates in the skeletal development directly or indirectly. Currently, the calcium agent and growth hormone-like drugs have provided potential opportunities to promote growth and development among children, yet limited trials existed with probiotic intake as a primary strategy.

**Objectives:** Present cohort study was designed to evaluate the height-promoting effect of gut probiotic *Bifidobacterium animalis subsp. lactis* BL-11 among 3-7 years old children. Registered under ClinicalTrials.gov Identifier no. ChiCTR2400089723.

**Methods:** This study was conducted with Chinese children aged 3 to 7 years (n = 124) who fall between the 3rd and 25th percentiles (P3-P25) for height, and whose annual growth rate in height is less than 5 cm. The *Bifidobacterium animalis subsp. lactis* BL-11 oiling agent were taken every day with total live bacteria ≥ 30 billion. The height of each participant was measured and recorded before and after three-months *Bifidobacterium animalis subsp. lactis* BL-11 intervention.

**Results:** *Bifidobacterium animalis subsp. lactis* BL-11 could strikingly promote the height growth of children, and shift the height percentile rage from P3-P10 to P10-P25. Importantly, this promoting effect has no specificity between age and sex. Comparatively, children with shorter height (P3-P10) were more sensitive to *Bifidobacterium animalis subsp. lactis* BL-11.

**Conclusions:** Administration of *Bifidobacterium animalis subsp. lactis* BL-11 as early as possible is in favor of the child’s growth and development, which provided a strategy for pediatricians to address some plights that how to promote children’s growth effectively and safely.

## Introduction

Recently, children suffering from growth delay have received social attentions, as the growth problem during early childhood higher the risk of long-term health, educational, and emotional problems for children [1,2]. According to World Health Organization (WHO) Global Database on Child Growth and Malnutrition, a child whose height is lower than the average age by more than two standard deviations is regarded as short stature [3]. Compelling evidences have proved that the short stature is closely related to the development of several disease, such as metabolic disease and cardiovascular diseases [4,5]. It is reported that an individual with short stature had about a 50% higher risk of coronary heart disease morbidity and mortality than tall ones [6]. Additionally, short stature can contribute to feelings of inferiority in children and negatively impact the development of a healthy personality. Hence, it is necessary to recognize the height problem in children as early as possible and to implement timely interventions.

An intestinal probiotic is a type of bacteria that has been found to be non-harmful and to have beneficial effects on the host [7-9]. Various studies have demonstrated that administering *Bifidobacterium* spp. can enhance human immunity and improve disease resistance [10]. For instance, *Bifidobacterium animalis subsp. Lactis* BB-12® has been shown to reduce crying and fussing in infants diagnosed with infant colic [11]. Additionally, treatment with *Lactobacillus acidophilus* DDS-1 and *Bifidobacterium animalis subsp. lactis* UABla-12 alleviates the abdominal pain in patients with irritable bowel syndrome [12]. *Bifidobacterium animalis subsp. lactis* Bb12 may alleviate the unhealthy conditions of preterm infants [13]. Moreover, a meta-analysis demonstrated that probiotic *Bifidobacterium animalis subsp. lactis* CNCM I-3446 could exert positive effect on growth in infants born to mothers with human immunodeficiency virus (HIV) [14]. Thus, probiotic intervention appears to be a promising strategy for alleviating disease and enhancing human health, particularly for children and patients who are intolerant to conventional medications. However, the effectiveness of probiotics in promoting children’s growth remains controversial and requires further clinical research to validate this hypothesis.

To explore the potential efficacy of probiotics in promoting children’s growth, we selected a strain of *Bifidobacterium animalis subsp. lactis* BL-11 which has shown to promote height in children with Prader-Willi Syndrome [15], and administered it to children aged 3-7 years for three months. The height were measured and recorded before and after intervention. As expected, there was a significant increase in height following the *Bifidobacterium animalis subsp. lactis* BL-11 intervention. This cohort study not only proved the role of probiotics in promoting children’s growth, but also provided an effective intervention for addressing short stature in children.

## Methods

### Subjects and ethical approval

Children of both sexes and aged 3-7 years who were recruited between Dec. 2023 and Sep. 2024. The study was conducted at the second affiliated hospital of anhui medical university. The enrollment and research plan were reviewed and approved by the institutional ethics committee of the second affiliated hospital of anhui medical university (ethics number: SL-YX2023-172). Written informed consent was obtained from each child’s parents/guardians. This study complied with the code of ethics of the World Medical Association (Declaration of Helsinki) and received approval and registration in the Chinese Clinical Trial Registration Center with the registration number ChiCTR2400089723.

### Inclusion, exclusion, and withdrawal criteria

#### Inclusion Criteria

1. Aged 3-7 years.
2. Height between the 3rd and 25th percentiles (P3∼P25),
3. The annual growth rate of height is less than 5 cm.

#### Exclusion Criteria

1. Inherited metabolic diseases: trisomy 21 syndrome, Turner syndrome, hypothyroidism, hyperthyroidism, growth hormone deficiency, SGA, hypophosphatemic anti-D rickets.
2. Use of various antibiotics, antimicrobials, immunosuppressive drugs, other probiotics, prebiotics, various dietary therapies, large doses of nutrients, and other concurrent alternative therapies within the past one month.
3. Bone metabolic diseases: achondroplasia, osteogenesis imperfecta, moderate to severe nutritional rickets.
4. Chronic diarrhea, asthma.
5. Other underlying diseases (leukemia, thalassemia, aplastic anemia, etc.).
6. Other circumstances considered by the investigator to be inappropriate for inclusion in the trial.

#### Withdrawal criteria

1. Misdiagnosis and incorrect admission.
2. Eligible patients who met the inclusion criteria did not take drugs after enrollment;
3. No post-treatment records.
4. During the trial, subjects used drugs that influenced the efficacy of the study drugs that were prohibited by the study.

#### Intervention

Dose, route and duration of administration

a. Take 40 drops of BL-11 daily (20 drops twice a day, recommended half an hour after breakfast and half an hour before going to bed for best results).
b. Shake thoroughly and then drop into the mouth or add to other food to eat.
c. Continue for 3 months.

Remark: Subjects or their guardians should properly use the study drugs under appropriate guidance and training. The precautions for drug use are as follows:

a. Take care that the mouth of the bottle and dropper do not touch the skin and saliva during the eating process.
b. Add oil droplets to foods or drinks at temperatures not higher than 37 °C, which can impair probiotic activity.
c. If the probiotic oil drops are dropped into water or liquid food for consumption, it is recommended to drink within 20 minutes in order to maintain the activity of probiotics.
d. Shake the product well before eating, shaking helps the probiotics to disperse more evenly.
e. Do not take it with antibiotics. If antibiotics are used, take it at least 2 hours apart. f, it is recommended to use it within 45 days after opening the lid.

At the same time, subjects or their guardians should carefully fill in the patient diary card to record the patient’s medication process.

#### Data collection

Height measurements: Height measurements were performed by the same, trained researchers as much as possible at the same time in the morning. Height was measured with the use of a calibrated wall height meter, and three values were measured and recorded at each visit. If the 3 measurements of height are different from each other. 0.5 cm, then the height was measured three times again until the difference between the two measurements was ≤0.5 cm.

To measure your height, use the following procedure:

Height measurements were recommended to be performed in the morning by the same trained researcher.

1. Use a calibrated height meter.
2. Subjects should not stretch before height determination.
3. The subjects had to be standing with no shoes (bare feet or socks only).
4. The subject should wear only light clothing so that the body posture of the subject can be observed.
5. Subjects had to look ahead horizontally.
6. Heel must be close together. If the subject had genu valgum (knee buckle), the knees had to touch each other and the heels had to be as close to each other as possible.
7. Heel, buttocks, shoulders and occipital bones must be in contact with the height measuring instrument.
8. The mandible (chin) must be tightened upward.
9. The shoulders should be relaxed and the abdomen tightened to reduce lordosis (curvature of the spine).
10. Lower the slide ruler or weight headrest until it is in contact with the highest part of the subject’s head.
11. The height measurement person read and recorded the measurement value.
12. Have the subject step away from the height meter and repeat two more times according to the previous procedure. Repeated measurements had to differ by 0.5 cm or less between the two measurements. If the 3 measurements of height are different from each other. 0.5 cm, then the height measurements were repeated three times until the difference between the two measurements was ≤0.5 cm.
13. Record the measurement value and measurement time.

### Statistics

All the information obtained was transferred to a Microsoft Excel® 2021 table where age in months, sex, height, and percentiles were recorded. GraphPad Prism 8 (San Diego, USA) was employed for statistical analysis. Data were presented as mean ± SEM, and figures were generated using the same software. The unpaired Student’s *t*-test was utilized to assess differences. Statistical significance was established at P < 0.05.

## Results

### Study design and characteristics of participants

To evaluate the growth-promoting effect of *Bifidobacterium animalis subsp. lactis* BL-11 (BL-11), we enrolled 124 participates, consisting of 60 boys and 64 girls. These children ranged in age from 3 to 7 years, with the average age of 60.8 months. The height of participants ranged from 92 to 117 cm, with a mean height of 104.6 cm **(Table 1)**. Each participant consumed 40 drops of BL-11 oiling agent daily (total live bacteria ≥ 30 billion/day) for 3 months. Heights were measured and recorded both before and after the BL-11 intervention. Finally, 124 participants with valid data were included in the height assessment. Detailed information is presented in **Supplementary Table 1**.

**Table 1.** Characteristics of the study population.

| Variables | Before intervention | After intervention |
| --- | --- | --- |
| Gender, male/female (n/n) | 60/64 | 60/64 |
| Age, mean (months) | 60.798 | 63.798 |
| Height, mean (cm) | 104.628 | 106.854 |

### Effect of Bifidobacterium animalis subsp. lactis BL-11 on height in pediatric subjects

We analyzed height changes among the 124 participants. On average, children who consumed BL-11 experienced a height increase of 2.23 cm over three months **(Table 1**, **Figure 1A)**. The included children were in the preschool stage, during which the typical annual height increase is 5-6 cm. Therefore, we examined the proportion of children whose height growth rate exceeded the average expected rate of 1.5 cm over three months. A height increase greater than 1.5 cm was observed in 104 children (accounting to 83.87%), while the remaining 20 children (accounting to 16.13%) showed an increase of 1.5 cm or less **(Figure 1B-C)**.

**Figure 1.**
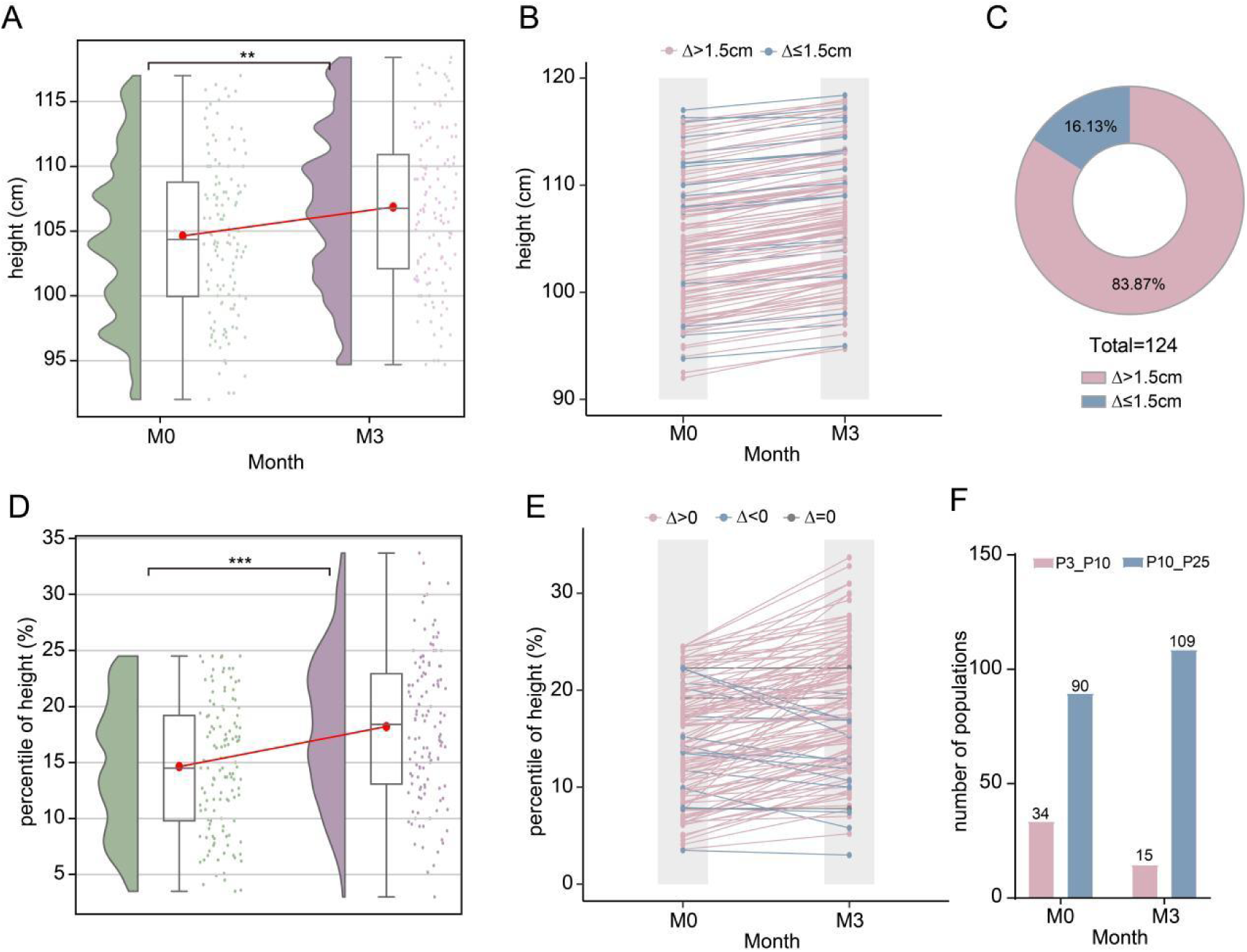
Effect of *Bifidobacterium animalis subsp. lactis* BL-11 on height in pediatric subjects. (A) The height of 124 children before and after 3-months BL-11 intervention. (B) The height change of 124 children. Red line represents a height increase greater than 1.5 cm; blue line represents a height increase less than 1.5 cm. (C) The proportions of children whose height has increased by more than 1.5 cm and less than 1.5 cm. (D) The height percentile of 124 children before and after 3-months BL-11 intervention. (E) The height percentile change of 124 children. Red line represents an increase; blue line represents a decline; gray line represents unchanged. (F) The number of children in P3-P10 and P10-P25 rages before and after 3-months BL-11 intervention. \*\**P*<0.01, \*\*\**P*<0.001.

The height percentile reflects the percentage of subjects with a certain height relative to the total number of subjects. We next analyzed height percentile before and after the BL-11 intervention. A significant increase in height percentile was observed post- intervention **(Figure 1D)**. Of the 124 children, 108 showed an increase in height percentile, 13 showed a decrease, and 3 showed no change **(Figure 1E)**.

To further understand the impact of BL-11, we categorized the into two groups based on height percentile: P3-P10 and P10-P25. Before the BL-11 treatment, 34 children were in the P3-P10 percentile range, and 90 children were geared to P10-P25 range. After BL-11 treatment, there was a notable shift, with only 15 children remaining in the P3-P10 range and 109 children moving to the P10-P25 range **(Figure 1F)**. These results suggested that BL-11 effectively promotes height growth in children, particularly those with shorter stature.

### Effect of Bifidobacterium animalis subsp. lactis BL-11 on height in children by gender

To determine if the growth-promoting effect of BL-11 varies by gender, we conducted a subgroup analysis of the cohort based on gender. The height tended to be increased among both male and female children after the three-month BL-11 intervention, although the increase was not statistically significant **(Figure 2A)**. Among the 60 male participants, 50 (83.33%) experienced a height increase greater than 1.5 cm, while 10 (16.67%) showed an increase of 1.5 cm or less **(Figure 2B-C)**. Similarly, of the 64 female participants, 54 (84.38%) had a height increase greater than 1.5 cm, and 10 (15.62%) had an increase of 1.5 cm or less **(Figure 2B-C)**.

**Figure 2.**
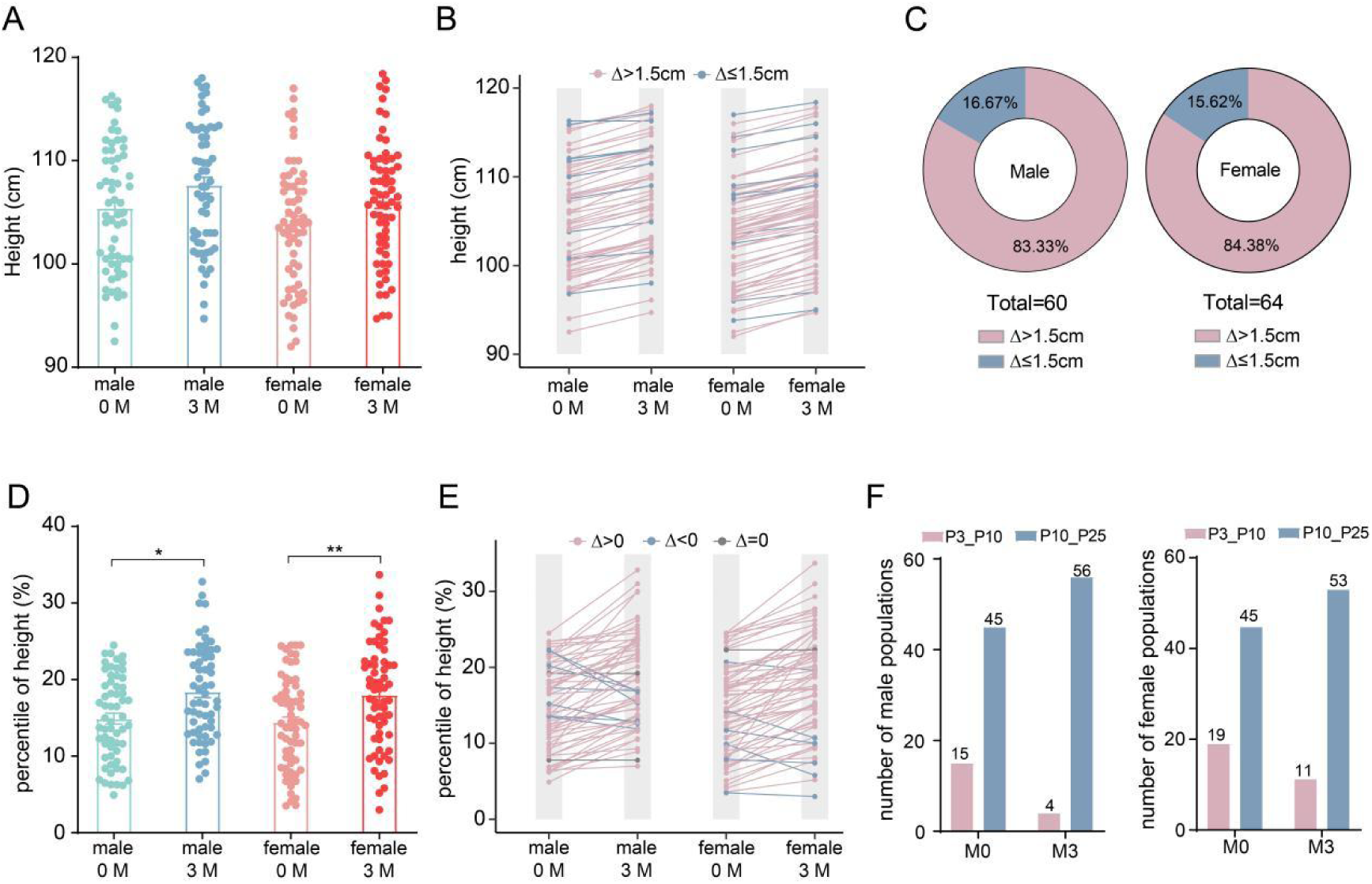
Effect of *Bifidobacterium animalis subsp. lactis* BL-11 on height in children by gender. (A) The height of male and female children before and after 3-months BL-11 intervention. (B) The height change of male and female children. Red line represents a height increase greater than 1.5 cm; blue line represents a height increase less than 1.5 cm. (C) The proportions of male and female children whose height has increased by more than 1.5 cm and less than 1.5 cm, respectively. (D) The height percentile of male and female children before and after 3-months BL-11 intervention. (E) The height percentile change of male and female children. Red line represents an increase; blue line represents a decline; gray line represents unchanged. (F) The number of male or female children in P3-P10 and P10-P25 rages before and after 3-months BL-11 intervention. \**P*<0.05, \*\**P*<0.01.

Although the overall height increase was not statistically significant, the height percentiles showed a notable improvement in both male and female children after BL-11 treatment **(Figure 2D)**. Among the male participants, 51 showed an increase in height percentile, 7 showed a decrease, and 2 showed no change **(Figure 2E)**. Among the female children, 57 showed an increase in height percentile, 6 showed a decrease, and 1 showed no change **(Figure 2E)**.

At the baseline, 15 male and 19 female children were in the P3-P10 percentile range, while 45 male and 45 female children were in the P10-P25 percentile range. Post the BL-11 intervention, only 4 male and 11 female children remained in the P3-P10 range, whereas 56 male and 53 female individuals moved to the P10-P25 range **(Figure 2F)**. Above data indicate that BL-11 effectively promotes height growth in children regardless of gender.

### Effect of Bifidobacterium animalis subsp. lactis BL-11 on height in children by age

Next, we analyzed the age distribution of our cohort, dividing it into six stages: 3 children aged 30-40 months, 24 children aged 40-50 months, 29 children aged 50-60 months, 40 children aged 60-70 months, 20 children aged 70-80 months, and 8 children aged 80-90 months. For further analysis, we divided the children into two subgroups based on 60 months as the boundary, with 56 children under 60 months old and 68 participates aged 60 months or older **(Figure 3A)**. Overall, the height was markedly increased in both two subgroups **(Figure 3B)**. Among the children under 60 months old, 50 individuals (89.29%) experienced a height increase greater than 1.5 cm, while 6 participants (10.71%) showed an increase of 1.5 cm or less **(Figure 3C-D)**. For those aged 60 months or older, 54 individuals (79.41%) had a height growth rate greater than 1.5 cm, while 14 individuals (20.59%) presented a height growth rate less than 1.5 cm **(Figure 3C,E)**.

**Figure 3.**
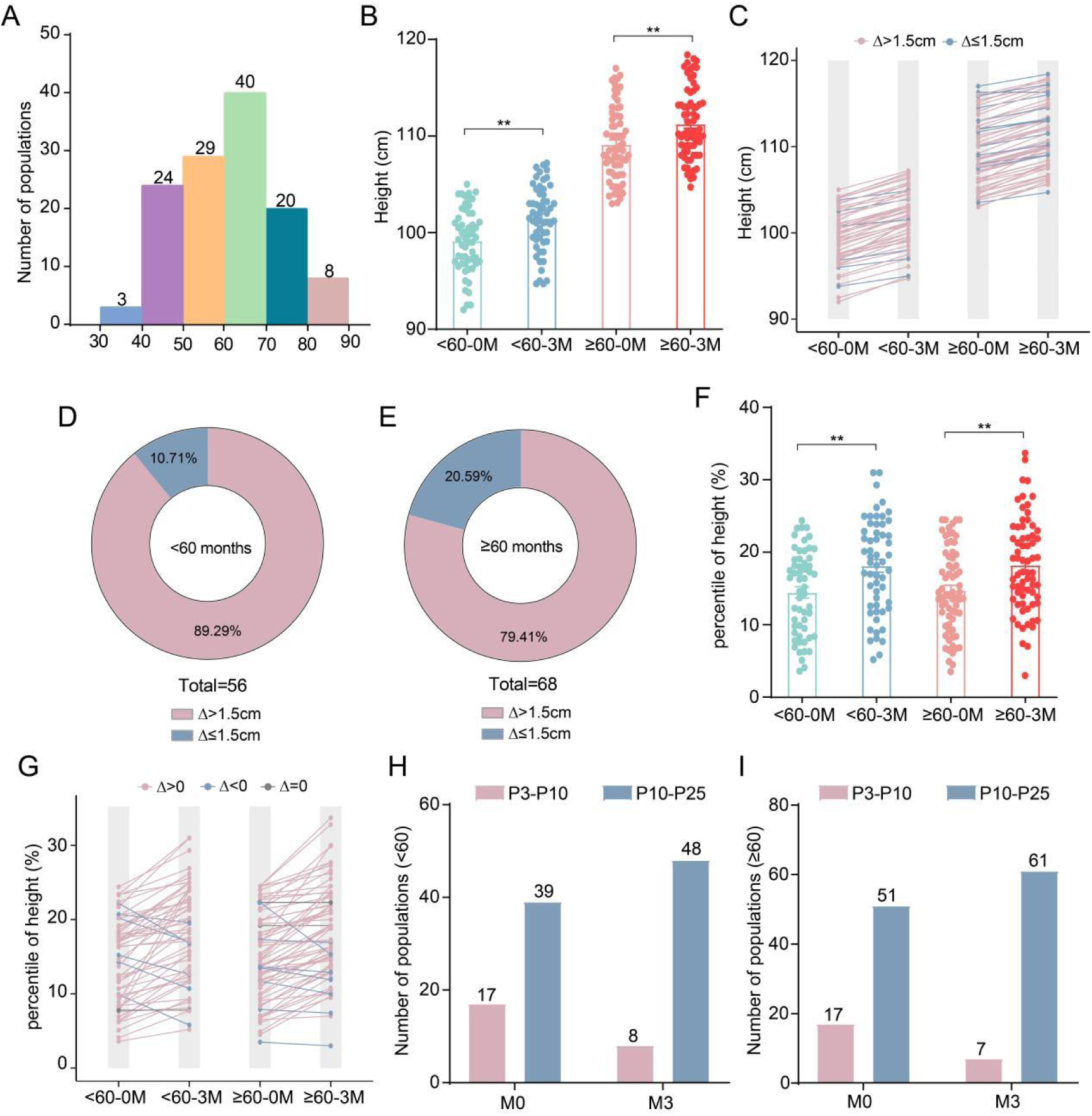
Effect of *Bifidobacterium animalis subsp. lactis* BL-11 on height in children by age. (A) The age distribution of 124 children. (B) The height of children aged under 60 months or over 60 months before and after 3-months BL-11 intervention. (C) The height change of children aged under 60 months or over 60 months. Red line represents a height increase greater than 1.5 cm; blue line represents a height increase less than 1.5 cm. (D) The proportions of children aged under 60 months whose height has increased by more than 1.5 cm and less than 1.5 cm. (E) The proportions of children aged over 60 months whose height has increased by more than 1.5 cm and less than 1.5 cm. (F) The height percentile of children aged under 60 months or over 60 months before and after 3-months BL-11 intervention. (G) The height percentile change of children aged under 60 months or over 60 months. Red line represents an increase; blue line represents a decline; gray line represents unchanged. (H) The number of children aged under 60 months in P3-P10 and P10-P25 rages before and after 3-months BL-11 intervention. (I) The number of children aged over 60 months in P3-P10 and P10-P25 rages before and after 3-months BL-11 intervention. \*\**P*<0.01.

The height percentile also strikingly increased in both subgroups **(Figure 3F)**. Specifically, among children under 60 months old, the percentile decreased in 6 children and increased in 49 children. Among children aged 60 months or older, the percentile decreased in 7 children and increased in 59 children **(Figure 3G)**.

At the baseline, 17 children in both age groups were in the P3-P10 percentile range, while 39 children children under 60 months old and 51 children aged 60 months 0r older were in the P10-P25 percentile range, Post the BL-11 intervention, only 8 children under 60 months old and 7 children aged 60 months or older remained in the P3-P10 range, whereas 48 children under 60 months old and 61 children aged 60 months or older moved to the P10-P25 range **(Figure 3F)**. These data indicate that BL-11 effectively enhances height growth in children regardless of age.

### Effect of Bifidobacterium animalis subsp. lactis BL-11 on height in children within different height ranges

Based on the above analyses, we found that BL-11 has different effects on the height of children within the P3-P10 and P10-P25 height percentiles. Therefore, we reanalyzed the population within the in these two height ranges. The height showed an increasing trend in both the P3-P10 and P10-P25 groups following the BL-11 intervention, although the increase was not statistically significant **(Figure 4A)**. Within the P3-P10 range, 29 (85.29%) children experienced a height increase greater than 1.5 cm, while 5 (14.71%) children increased less than 1.5 cm **(Figure 4B-C)**. Comparatively, in the P10-P25 range, 75 (83.33%) children had a height increase greater than 1.5 cm, while 15 (16.67%) children had an increase less than 1.5 cm **(Figure 4B-C)**.

**Figure 4.**
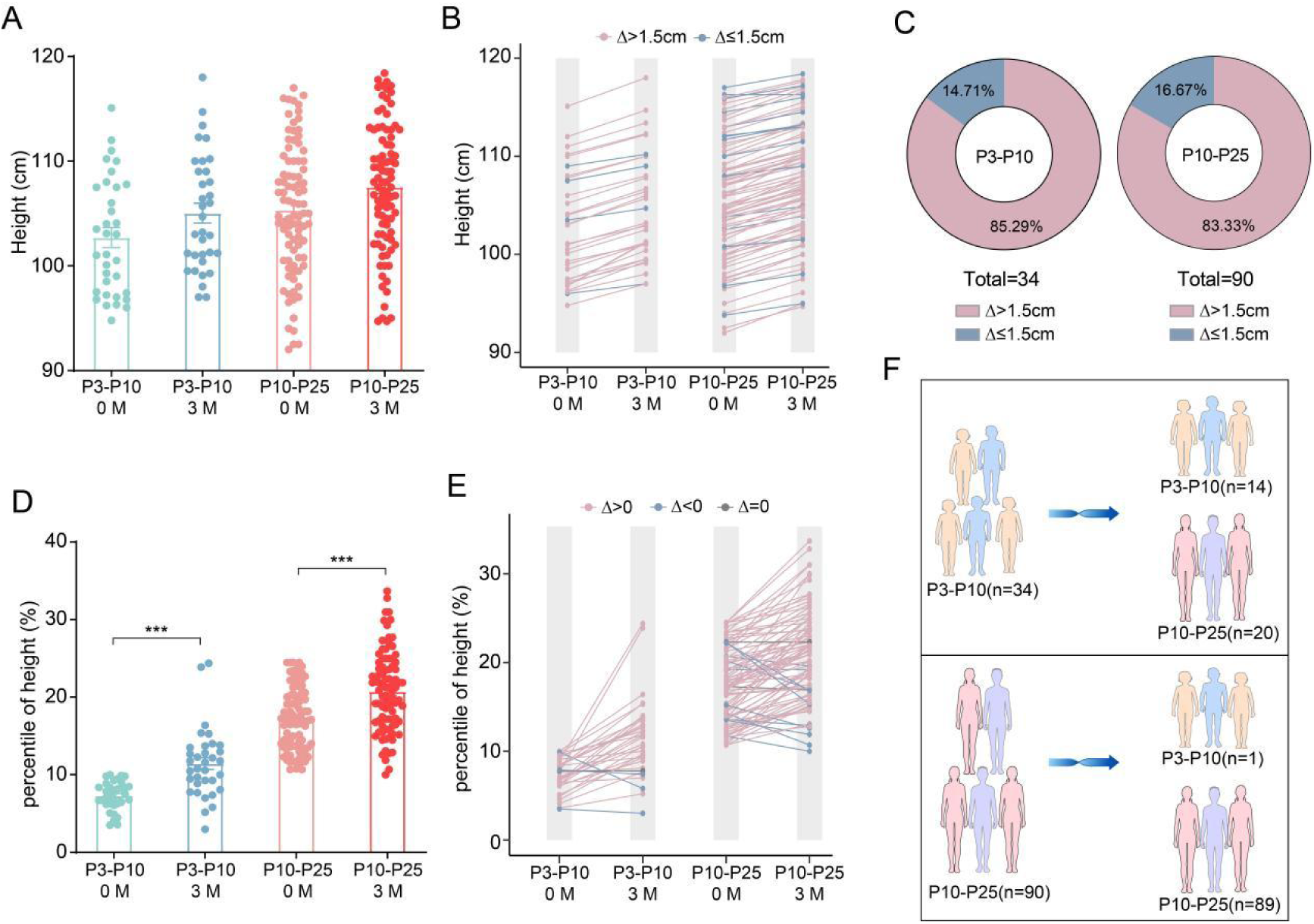
Effect of *Bifidobacterium animalis subsp. lactis* BL-11 on height in children within different height ranges. (A) The height of children within the P3-P10 and P10-P25 ranges before and after 3-months BL-11 intervention. (B) The height change of children within the P3-P10 and P10-P25 ranges. Red line represents a height increase greater than 1.5 cm; blue line represents a height increase less than 1.5 cm. (C) The proportions of children within the P3-P10 and P10-P25 ranges whose height has increased by more than 1.5 cm and less than 1.5 cm, respectively. (D) The height percentile of children within the P3-P10 and P10-P25 ranges before and after 3-months BL-11 intervention. (E) The height percentile change of children within the P3-P10 and P10-P25 ranges. Red line represents an increase; blue line represents a decline; gray line represents unchanged. (F) The number of children in P3-P10 and P10-P25 rages before and after 3-months BL-11 intervention. \*\*\**P*<0.001.

The height percentile notably increased in both the P3-P10 and P10-P25 groups following the BL-11 intervention **(Figure 4D)**. Among children in the P3-P10 range, 30 showed an increase in height percentile, while 3 showed a decrease **(Figure 4E)**. In the P10-P25 range, 78 children showed an increase inn height percentile, while 10 children showed a decline **(Figure 4E)**. Overall, BL-11 effectively shifted the height range of 20 children from P3-P10 to P10-P25 **(Figure 4F)**, displaying a superior growth-promoting effect.

## Discussions

Early childhood is a period of rapid development in metabolism, endocrine, immunity, and neural development, all of which strongly affect the growth and development of children. Therefore, it is necessary to explore strategies to promote the growth of children with short stature. The present cohort assessed the growth-promoting effect of *Bifidobacterium animalis subsp. lactis* BL-11 among 124 children aged 3-7 years old with height ranging from 92 to 117. Positively, three-months intervention with *Bifidobacterium animalis subsp. lactis* BL-11 significantly promoted height growth in these children, with no difference observed based on gender or age. To a certain extent, these clinical trial data demonstrate that treatment with specific probiotics strain can favorably impact children’s growth and development. This finding is likely to attract the attention of pediatrician and parents concerned about short stature in children.

The chief indicators of short stature in children are their height and weight. Growth disorders, such as short stature or growth failure, are often markers of underlying conditions like nutritional deficits, abnormal hormone metabolism, diarrhoeal disease or cardiovascular disease [16]. Currently, measures to promote children’s growth primarily include calcium agent supplementation and growth hormone-like drugs. While these treatments are effective for many individuals, the daily injections can be painful and and intolerable, particularly for children, leading to the non-adherence and reduced treatment outcomes [17]. Obviously, it is meaningful to explore complementary or alternative therapies that are both effective and safe.

The gut microbiota, which colonizes and stabilizes within the human intestine during the childhood, plays a crucial role in growth and development. Compelling evidences have confirmed that those gut bacteria contribute to lifelong and intergenerational deficits in growth and development among children [18]. Probiotic strains of *Bifidobacterium* are increasingly being used in pharmaceuticals and functional foods [19]. *Bifidobacterium animalis subsp. lactis* is one of the probiotics that supports the intestinal health, metabolic balance and growth performance [20-22], indicating its potential as a candidate for height management. Consistent with the speculation, our study found that *Bifidobacterium animalis subsp. lactis* BL-11 notably enhanced the height of children, implying that early probiotic supplementation might prevent the occurrence of short status.

Generally, children’s height is determined by their longitudinal growth index, which is influenced by hereditary factors, nutrition, sleep, sports, hormone level, environment, habits, and race [23]. Several *Bifidobacterium* strains, such as *Bifidobacterium longum* and *Bifidobacterium pseudocatenulatum* CECT 7765, are demonstrated to increase serum osteocalcin and minerals concentration, along with decrease serum C-terminal telopeptide and parathormone, thereby favor a femur growth [24,25]. Our present study displayed growth-promoting effect of another *Bifidobacterium* strain — *Bifidobacterium animalis subsp. lactis* BL-11. While the study was promising, it failed to provide mechanisms since mechanistic studies were not performed. Ding et al. revealed that strain of *Bifidobacterium longum subsp. infantis* CCFM1269 can modulate gut microbiota and metabolites, stimulate GH/IGF-1 axis in growing mice [26]. We, therefore, cautiously speculate that the growth-promoting mechanism of *Bifidobacterium animalis subsp. lactis* BL-11 is related with the GH/IGF-1 pathway. Additionally, it is reported that Bifidobacterium longum can modulate Treg-Th17 cell balance, elevate anti-osteoclastogenic cytokines IFN-γ and IL-10 and decrease osteoclastogenic-cytokines IL-6, IL-17, and TNF-α [27]. Hence, the immunomodulatory potential of *Bifidobacterium animalis subsp. lactis* BL-11 may also be conducive to its growth-promoting effect. A further study should elucidate the mechanisms of how *Bifidobacterium animalis subsp. lactis* BL-11 promote bone growth, thus promote effective and efficient probiotic products development.

## Conclusions

Collectively, the foregoing data demonstrated that *Bifidobacterium animalis subsp. lactis* BL-11 intervention is in favor of the child’s growth and development. This cohort study proposes that early supplementary with *Bifidobacterium animalis subsp. lactis* BL-11 could help prevent short stature in children. These findings provide pediatricians with a potential strategy to effectively and safely promote children’s growth.

## Author Approval

All authors have seen and approved the manuscript.

## Acknowledgments

None

## CRediT authorship contribution statement

Q.W.: Methodology, Formal Analysis, Visualization, Writing-Review & Editing; Y.Y.: Methodology, Formal Analysis, Visualization, Writing-Review & Editing; Y.X.: Visualization, Formal Analysis, Writing-original draft; Z.L.: Visualization, Formal Analysis, Writing-original draft; M.L.: Methodology, Writing-Review & Editing; Y.Y.: Methodology, Writing-Review & Editing; N.W.: Methodology, Investigation; J.L.: Methodology, Investigation; X.L.: Visualization; C.X.: Visualization; D.L.: Conceptualization, Data Curation, Formal Analysis, Visualization, Writing-Review & Editing; C.W.: Conceptualization, Data Curation, Formal Analysis, Visualization, Writing-Review & Editing.

## Data availability

All data are available from the corresponding authors upon reasonable request.

